# Large language models approach expert-level clinical knowledge and reasoning in ophthalmology: A head-to-head cross-sectional study

**DOI:** 10.1101/2023.07.31.23293474

**Authors:** Arun James Thirunavukarasu, Shathar Mahmood, Andrew Malem, William Paul Foster, Rohan Sanghera, Refaat Hassan, Sean Zhou, Shiao Wei Wong, Yee Ling Wong, Yu Jeat Chong, Abdullah Shakeel, Yin-Hsi Chang, Benjamin Kye Jyn Tan, Nikhil Jain, Ting Fang Tan, Saaeha Rauz, Daniel Shu Wei Ting, Darren Shu Jeng Ting

## Abstract

**Objective:** To evaluate the clinical potential of large language models (LLMs) in ophthalmology using a more robust benchmark than raw examination scores.

**Materials and methods:** GPT-3.5 and GPT-4 were trialled on 347 questions before GPT-3.5, GPT-4, PaLM 2, LLaMA, expert ophthalmologists, and doctors in training were trialled on a mock examination of 87 questions. Performance was analysed with respect to question subject and type (first order recall and higher order reasoning). Masked ophthalmologists graded the accuracy, relevance, and overall preference of GPT-3.5 and GPT-4 responses to the same questions.

**Results:** The performance of GPT-4 (69%) was superior to GPT-3.5 (48%), LLaMA (32%), and PaLM 2 (56%). GPT-4 compared favourably with expert ophthalmologists (median 76%, range 64-90%), ophthalmology trainees (median 59%, range 57-63%), and unspecialised junior doctors (median 43%, range 41-44%). Low agreement between LLMs and doctors reflected idiosyncratic differences in knowledge and reasoning with overall consistency across subjects and types (*p*>0.05). All ophthalmologists preferred GPT-4 responses over GPT-3.5 and rated the accuracy and relevance of GPT-4 as higher (*p*<0.05).

**Discussion:** In view of the comparable or superior performance to trainee-grade ophthalmologists and unspecialised junior doctors, state-of-the-art LLMs such as GPT-4 may provide useful medical advice and assistance where access to expert ophthalmologists is limited. Clinical benchmarks provide useful assays of LLM capabilities in healthcare before clinical trials can be designed and conducted.

**Conclusion:** LLMs are approaching expert-level knowledge and reasoning skills in ophthalmology. Further research is required to develop and validate clinical applications to improve eye health outcomes.

## INTRODUCTION

Generative Pre-trained Transformer 3.5 (GPT-3.5) and 4 (GPT-4) are large language models (LLMs) trained on datasets containing hundreds of billions of words from articles, books, and other internet sources.^1,2^ ChatGPT is an online chatbot which uses GPT-3.5 or GPT-4 to provide bespoke responses to human users’ queries.^3^ LLMs have revolutionised the field of natural language processing, and in recent months, ChatGPT has attracted significant attention in medicine for attaining passing level performance in medical school examinations and providing more accurate and empathetic messages than human doctors in response to patient queries on a social media platform.^3–6^ While GPT-3.5 performance in more specialised examinations has been inadequate, GPT-4 is thought to represent a significant advancement in terms of medical knowledge and reasoning.^3,7,8^ Other LLMs in wide use include Pathways Language Model 2 (PaLM 2) and Large Language Model Meta AI 2 (LLaMA 2).^3,9,10^

Applications and trials of LLMs in ophthalmological settings has been limited despite ChatGPT’s performance in questions relating to ‘eyes and vision’ being superior to other subjects in an examination for general practitioners.^7,11^ ChatGPT has been trialled on the North American Ophthalmology Knowledge Assessment Program (OKAP), and Fellowship of the Royal College of Ophthalmologists (FRCOphth) Part 1 and Part 2 examinations. In both cases, relatively poor results have been reported for GPT-3.5, with significant improvement exhibited by GPT-4.^12–16^ However, previous studies are afflicted by two important issues which may affect their validity and interpretability. First, so-called ‘contamination’, where test material features in the pretraining data used to develop LLMs, may result in inflated performance as models recall previously seen text rather than using clinical reasoning to provide an answer. Second, examination performance in and of itself provides little information regarding the potential of models to contribute to clinical practice as a medical-assistance tool.^3^ Clinical benchmarks are required to understanding the meaning and implications of scores in ophthalmological examinations attained by LLMs and are a necessary precursor to clinical trials of LLM-based interventions.

Here, we used FRCOphth Part 2 examination questions to gauge the ophthalmological knowledge base and reasoning capability of LLMs using fully qualified and currently training ophthalmologists as clinical benchmarks. These questions were not freely available online, minimising the risk of contamination. The FRCOphth Part 2 Written Examination tests the clinical knowledge and skills of ophthalmologists in training using multiple choice questions with no negative marking and must be passed to fully qualify as a specialist eye doctor in the United Kingdom.

## METHODS

### Question extraction

FRCOphth Part 2 questions were sourced from a textbook for doctors preparing to take the examination.^17^ This textbook is not freely available on the internet, making the possibility of its content being included in LLMs’ training datasets unlikely.^1^ All 360 questions from the textbook’s six chapters were extracted. Two researchers matched the subject categories of the practice papers’ questions to those defined in the Royal College of Ophthalmologists’ documentation concerning the FRCOphth Part 2 written examination. Similarly, two researchers categorised each question as first order recall or higher order reasoning, corresponding to ‘remembering’ and ‘applying’ or ‘analysing’ in Bloom’s taxonomy, respectively.^18^ Disagreement between classification decisions was resolved by a third researcher casting a deciding vote. Questions containing non-plain text elements such as images were excluded as these could not be inputted to the LLM applications.

### Trialling large language models

Every eligible question was inputted into ChatGPT (GPT-3.5 and GPT-4 versions; OpenAI, San Francisco, California, United States of America) between April 29 and May 10, 2023. The answers provided by GPT-3.5 and GPT-4 were recorded and their whole reply to each question was recorded for further analysis. If ChatGPT failed to provide a definitive answer, the question was re-trialled up to three times, after which ChatGPT’s answer was recorded as ‘null’ if no answer was provided. Correct answers (‘ground truth’) were defined as the answers provided by the textbook and were recorded for every eligible question to facilitate calculation of performance. Upon their release, Bard (Google LLC, Mountain View, California, USA) and HuggingChat (Hugging Face, Inc., New York City, USA) were used to trial PaLM 2 (Google LLC) and LLaMA (Meta, Menlo Park, California, USA) respectively on the portion of the textbook corresponding to a 90-question examination, adhering to the same procedures between June 20 and July 2, 2023.

### Clinical benchmarks

To gauge the performance, accuracy, and relevance of LLM outputs, five expert ophthalmologists who had all passed the FRCOphth Part 2 (E1-E5), three trainees (residents) currently in ophthalmology training programmes (T1-T3), and two unspecialised (*i.e.* not in ophthalmology training) junior doctors (J1-J2) first answered the 90-question mock examination independently, without reference to textbooks, the internet, or LLMs’ recorded answers. As with the LLMs, doctors’ performance was calculated with reference to the correct answers provided by the textbook. After completing the examination, ophthalmologists graded the whole output of GPT-3.5 and GPT-4 on a Likert scale from 1-5 (very bad, bad, neutral, good, very good) to qualitatively appraise accuracy of information provided and relevance of outputs to the question used as an input prompt. For these appraisals, ophthalmologists were blind to the LLM source (which was presented in a randomised order) and to their previous answers to the same questions, but they could refer to the question text and correct answer and explanation provided by the textbook. Procedures are comprehensively described in the protocol issued to the ophthalmologists (Supplementary Material 2).

As our null hypothesis was that LLMs and doctors would exhibit similar performance, prospective power analysis was conducted which indicated that 63 questions were required to identify a 10% superior performance of an LLM to human performance at a 5% significance level (type 1 error rate) with 80% power (20% type 2 error rate). This indicated that the 90-question examination in our experiments was more than sufficient to detect ∼10% differences in overall performance. The whole 90-question mock examination was used to avoid over- or under-sampling certain question types with respect to actual FRCOphth papers. To verify that the mock examination was representative of the FRCOphth Part 2 examination, expert ophthalmologists were asked to rate the difficulty of questions used here in comparison to official examinations on a 5-point Likert scale (“much easier”, “somewhat easier”, “similar”, “somewhat more difficult”, “much more difficult”).

### Statistical analysis

Performance of doctors and LLMs were compared using chi-squared (χ^2^) tests. Agreement between answers provided by doctors and LLMs was quantified through calculation of Kappa statistics, interpreted in accordance with McHugh’s recommendations.^19^ To further explore the strengths and weaknesses of the answer providers, performance was stratified by question type (first order fact recall or higher order reasoning) and subject using a chi-squared or Fisher’s exact test where appropriate. Likert scale data corresponding to the accuracy and relevance of GPT-3.5 and GPT-4 responses to the same questions were analysed with paired *t*-tests with the Bonferroni correction applied to mitigate the risk of false positive results due to multiple-testing—parametric testing was justified by a sufficient sample size.^20^ Statistical significance was concluded where *p* < 0.05. For additional contextualisation, examination statistics corresponding to FRCOphth Part 2 written examinations taken between July 2017 and December 2022 were collected from Royal College of Ophthalmologists examiners’ reports.^21^ These statistics facilitated comparisons between human and LLM performance in the mock examination with the performance of actual candidates in recent examinations.

Statistical analysis was conducted in R (version 4.1.2; R Foundation for Statistical Computing, Vienna, Austria), and figures were produced in Affinity Designer (version 1.10.6; Serif Ltd, West Bridgford, Nottinghamshire, United Kingdom).

## RESULTS

### Questions sources

Of 360 questions in the textbook, 347 questions (including 87 of the 90 questions from the mock examination chapter) were included.^17^ Exclusions were all due to non-text elements such as images and tables which could not be inputted into LLM chatbot interfaces. The distribution of question types and subjects within the whole set and mock examination set of questions is summarised in Table 1 and Table S1 alongside performance.

#### GPT-4 represents a significant advance on GPT-3.5 in ophthalmological knowledge and reasoning

Overall performance over 347 questions was significantly higher for GPT-4 (61.7%) than GPT-3.5 (48.41%; χ^2^=12.32, *p*<0.01), with results detailed in Figure S1 and Table S1. ChatGPT performance was consistent across question types and subjects (Table S1). For GPT-4, no significant variation was observed with respect to first order and higher order questions (χ^2^ = 0.22, *p*=0.64), or subjects defined by the Royal College of Ophthalmologists (Fisher’s exact test over 2000 iterations, *p* = 0.23). Similar results were observed for GPT-3.5 with respect to first and second order questions (χ^2^ = 0.08, *p* = 0.77), and subjects (Fisher’s exact test over 2000 iterations, *p* = 0.28). Performance and variation within the 87-question mock examination was very similar to the overall performance over 347 questions, and subsequent experiments were therefore restricted to that representative set of questions.

#### GPT-4 compares well with other LLMs, junior and trainee doctors and ophthalmology experts

Performance in the mock examination is summarised in Figure 1—GPT-4 (69%) was the top-scoring model, performing to a significantly higher standard than GPT-3.5 (48%; χ^2^ = 7.33, *p* < 0.01) and LLaMA (32%; χ^2^ = 22.77, *p* < 0.01), but statistically similarly to PaLM 2 (56%) despite a superior score (χ^2^ = 2.81, *p* = 0.09). LLaMA exhibited the lowest examination score, significantly weaker than GPT-3.5 (χ^2^ = 4.58, *p* = 0.03) and PaLM-2 (χ^2^ = 10.01, *p* < 0.01) as well as GPT-4.

**Figure 1:**
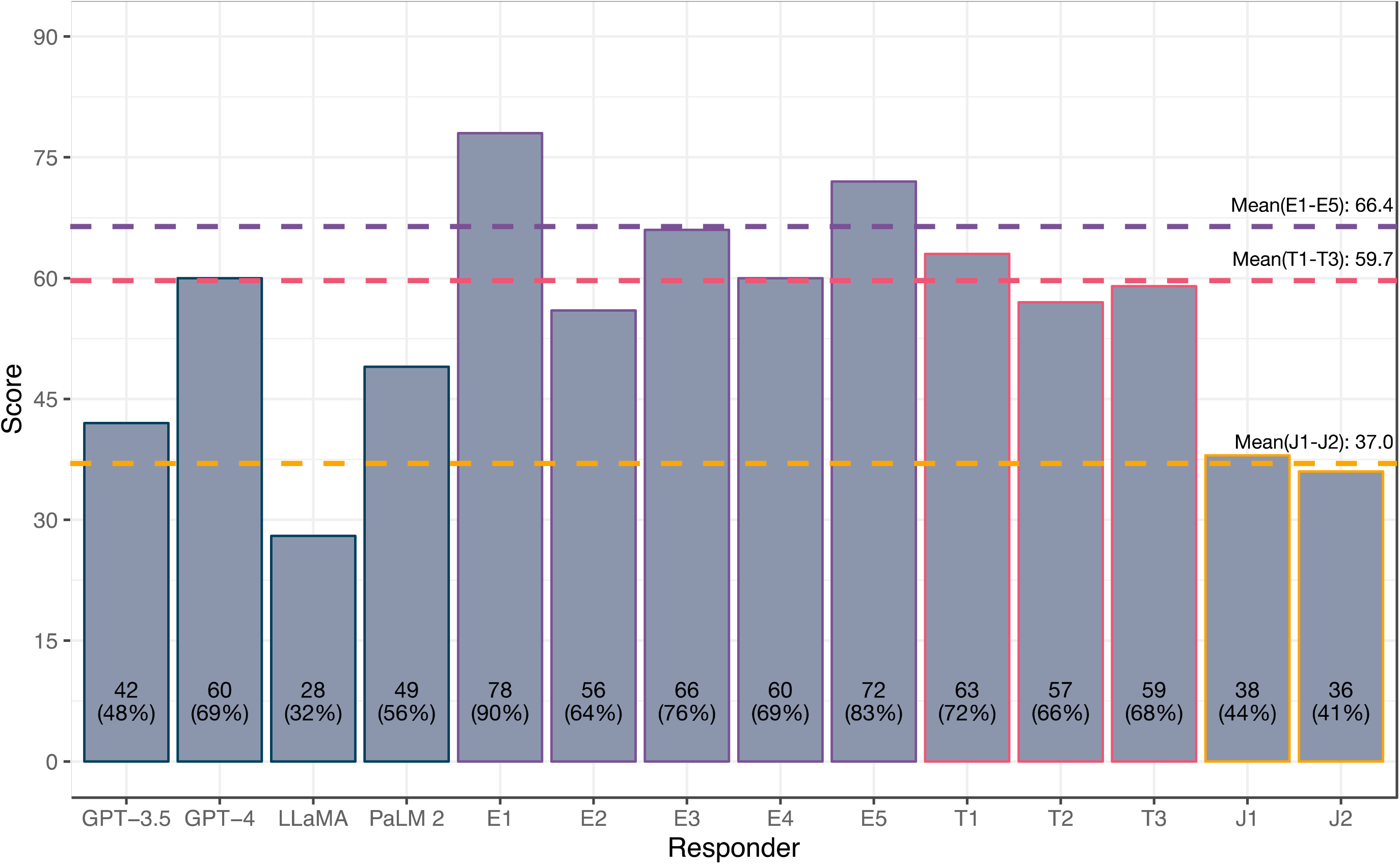
FRCOphth Part 2 performance of LLMs and doctors of variable expertise. Examination performance in the 87-question mock examination used to trial LLMs (GPT-3.5, GPT-4, LLaMA, and PaLM 2), expert ophthalmologists (E1-E5), ophthalmology trainees (T1-T3), and unspecialised junior doctors (J1-J2). Dotted lines depict the mean performance of expert ophthalmologists (66/87; 76%), ophthalmology trainees (60/87; 69%), and unspecialised junior doctors (37/87; 43%). The performance of GPT-4 lay within the range of expert ophthalmologists and ophthalmology trainees.

The performance of GPT-4 was statistically similar to the mean score attained by expert ophthalmologists (Figure 1; χ^2^ = 1.18, *p* = 0.28). Moreover, GPT-4’s performance exceeded the mean mark attained across FRCOphth Part 2 written examination candidates between 2017-2022 (66.06%), mean pass mark according to standard setting (61.31%), and the mean official mark required to pass the examination after adjustment (63.75%), as detailed in Table S2. In individual comparisons with expert ophthalmologists, GPT-4 was equivalent in 3 cases (χ^2^ tests, *p* > 0.05’ Table S3), and inferior in 2 cases (χ^2^ tests, *p* < 0.05; Table 2). In comparisons with ophthalmology trainees, GPT-4 was equivalent to all three ophthalmology trainees (χ^2^ tests, *p* > 0.05; Table 2). GPT-4 was significantly superior to both unspecialised trainee doctors (χ^2^ tests, *p* < 0.05; Table 2). Doctors were anonymised in analysis, but their ophthalmological experience is summarised in Table S3. Unsurprisingly, junior doctors (J1-J2) attained lower scores than expert ophthalmologists (E1-E5; *t* = 7.18, *p* < 0.01), and ophthalmology trainees (T1-T3; *t* = 11.18, *p* < 0.01), illustrated in Figure 1. Ophthalmology trainees approached expert-level scores with no significant difference between the groups (*t* = 1.55, *p* = 0.18). None of the other LLMs matched any of the expert ophthalmologists, mean mark of real examination candidates, or FRCOphth Part 2 pass mark.

Expert ophthalmologists agreed that the mock examination was a faithful representation of actual FRCOphth Part 2 Written Examination papers with a mean and median score of 3/5 (range 2-4/5).

#### LLM strengths and weaknesses are similar to doctors

Agreement between answers given by LLMs, expert ophthalmologists, and trainee doctors was generally absent (0 ≤ κ < 0.2), minimal (0.2 ≤ κ < 0.4), or weak (0.4 ≤ κ < 0.6), with moderate agreement only recorded for one pairing between the two highest performing ophthalmologists (Figure 2; κ = 0.64).^19^ Disagreement was primarily the result of general differences in knowledge and reasoning ability, illustrated by strong negative correlation between Kappa statistic (quantifying agreement) and difference in examination performance (Pearson’s r = -0.63, *p* < 0.01). Answer providers with more similar scores exhibited greater agreement overall irrespective of their category (LLM, expert ophthalmologist, ophthalmology trainee, or junior doctor).

**Figure 2:**
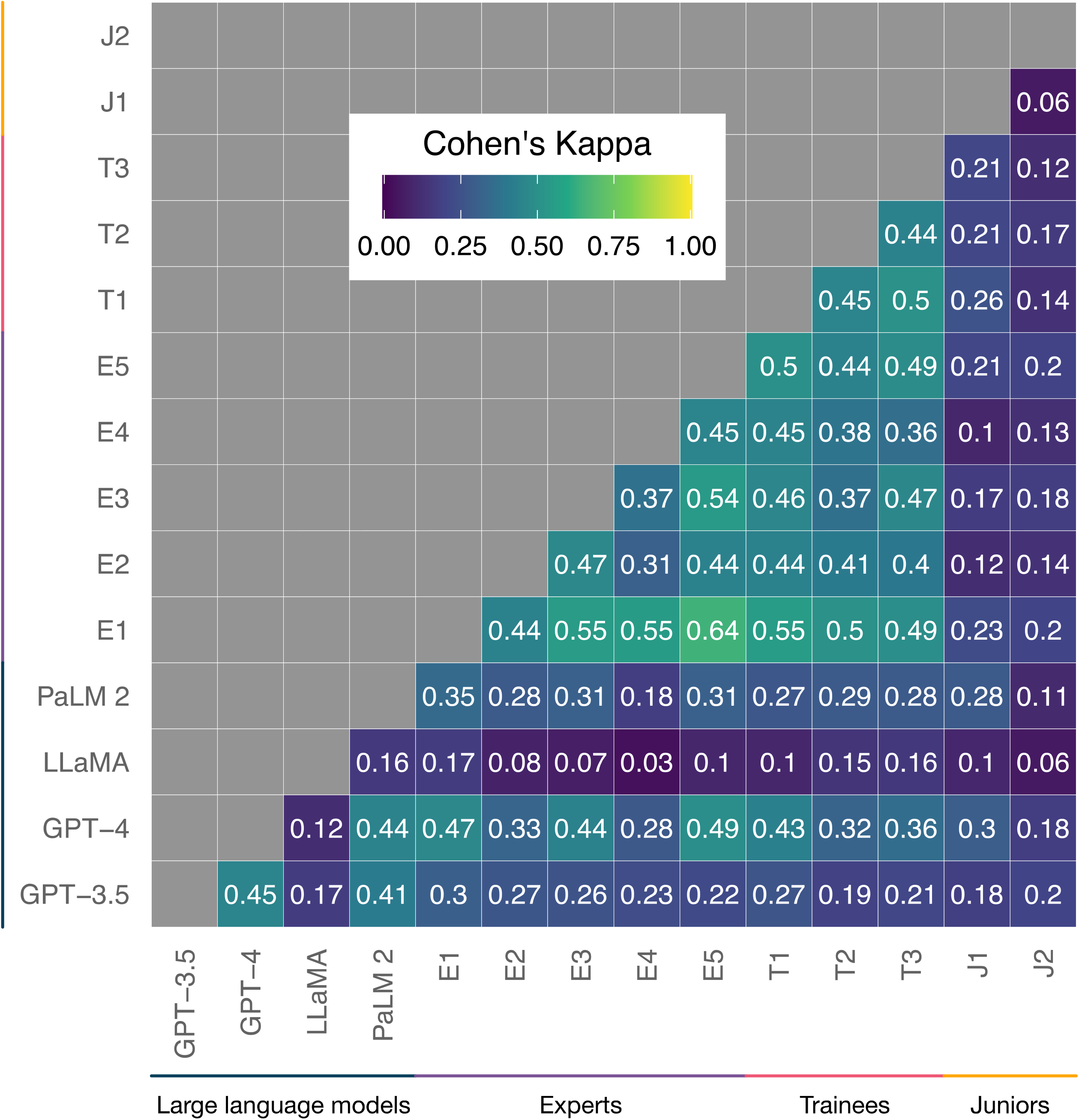
Heat map of Kappa statistics quantifying agreement between answers given by LLMs, expert ophthalmologists, and trainee doctors. Agreement correlates strongly with overall performance and stratification analysis found no particular question type or subject was associated with better performance of LLMs or doctors, indicating that LLM knowledge and reasoning ability is general across ophthalmology rather than restricted to particular subspecialties or question types.

Stratification analysis was undertaken to identify any specific strengths and weaknesses of LLMs with respect to expert ophthalmologists and trainee doctors (Table 1, Table S4). No significant difference between performance in first order fact recall and higher order reasoning questions was observed among any of the LLMs, expert ophthalmologists, ophthalmology trainees, or unspecialised junior doctors (Table S4; χ^2^ tests, *p* > 0.05). Similarly, only J1 (junior doctor yet to commence ophthalmology training) exhibited statistically significant variation in performance between subjects (Table S4; Fisher’s exact tests over 2000 iterations, *p* = 0.02); all other doctors and LLMs exhibited no significant variation (Fisher’s exact tests over 2000 iterations, *p* > 0.05). To explore whether consistency was due to an insufficient sample size, similar analyses were run for GPT-3.5 and GPT-4 performance over the larger set of 347 questions (Table S1; Table S4). As with the mock examination, no significant differences in performance across question types (Table S4; χ^2^ tests, *p* > 0.05) or subjects (Table S4; Fisher’s exact tests over 2000 iterations, *p* > 0.05) were observed.

#### LLM examination performance translates to subjective preference indicated by expert ophthalmologists

Ophthalmologists’ appraisal of GPT-4 and GPT-3.5 outputs indicated a marked preference for the former over the latter, mirroring objective performance in the mock examination and over the whole textbook. GPT-4 exhibited significantly (*t*-test with Bonferroni correction, *p* < 0.05) higher accuracy and relevance than GPT-3.5 according to all five ophthalmologists’ grading (Table 3). Differences were visually obvious, with GPT-4 exhibiting much higher rates of attaining the highest scores for accuracy and relevance than GPT-3.5 (Figure 3). This superiority was reflected in ophthalmologists’ qualitative preference indications: GPT-4 responses were preferred to GPT-3.5 responses by every ophthalmologist with statistically significant skew in favour of GPT-4 (χ^2^ test, *p* < 0.05; Table 3).

**Figure 3:**
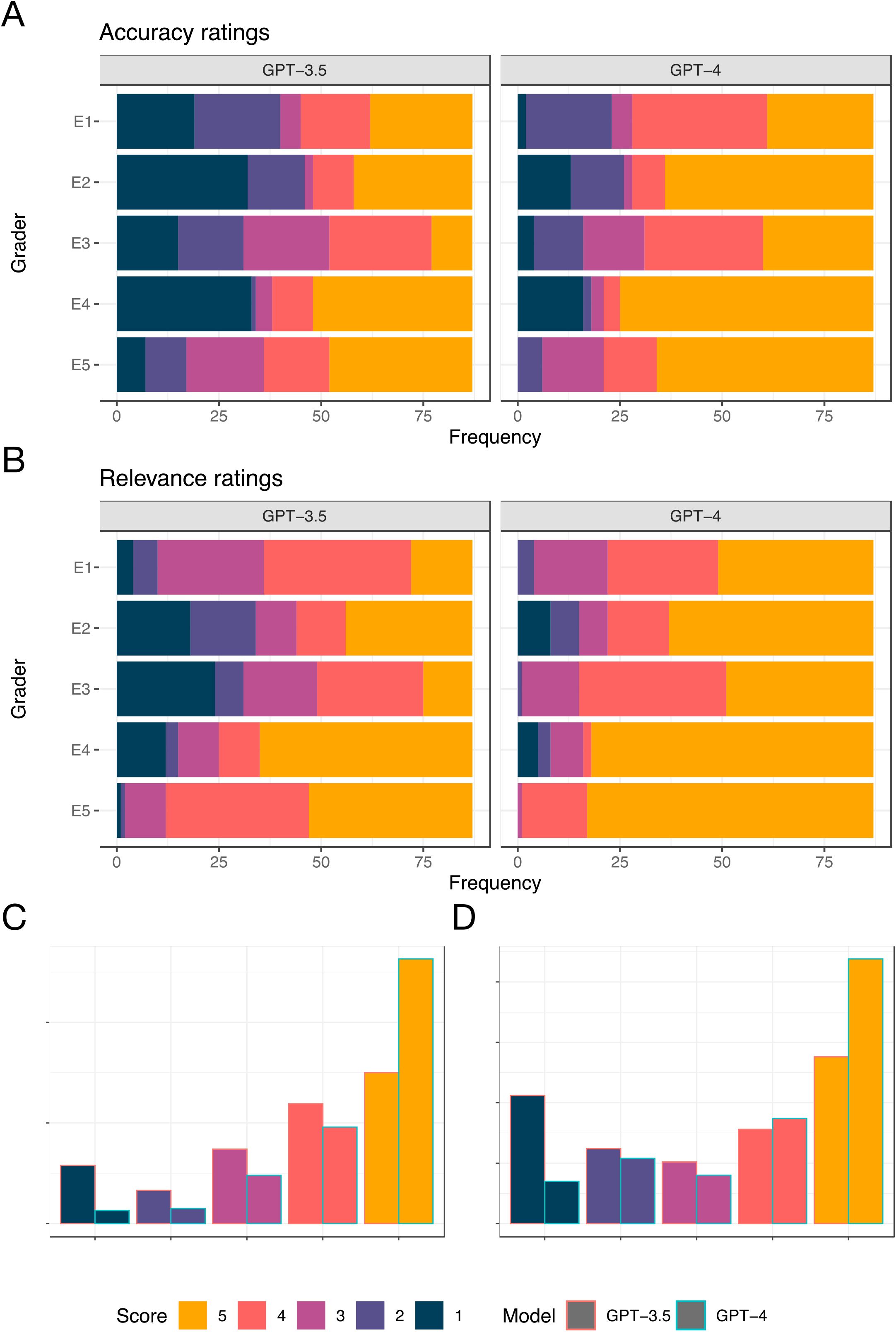
Accuracy and relevance of GPT-3.5 and GPT-4 in response to ophthalmological questions. Accuracy (A) and relevance (B) ratings were provided by five expert ophthalmologists for ChatGPT (powered by GPT-3.5 and GPT-4) responses to 87 FRCOphth Part 2 mock examination questions. In every case, the accuracy and relevance of GPT-4 is significantly superior to GPT-3.5 (t-test with Bonferroni correct applied, p < 0.05). Pooled scores for accuracy (C) and relevance (D) from all five raters are presented in the bottom two plots, with GPT-3.5 (left bars) compared directly with GPT-4 (right bars).

## DISCUSSION

Here, we present a clinical benchmark to gauge the ophthalmological performance of LLMs, using a source of questions with very low risk of contamination as the utilised textbook is not freely available online.^17^ Previous studies have suggested that ChatGPT can provide useful responses to ophthalmological queries, but often use online question sources which may have featured in LLMs’ pretraining datasets.^7,12,15,22^ In addition, our employment of multiple LLMs as well as fully qualified and training doctors provides novel insight into the potential and limitations of state-of-the-art LLMs through head-to-head comparisons which provide clinical context and quantitative benchmarks of competence in ophthalmology. Subsequent research may leverage our questions and results to gauge the performance of new LLMs and applications as they emerge.

We make three primary observations. First, performance of GPT-4 compares well to expert ophthalmologists and ophthalmology trainees, and exhibits pass-worthy performance in an FRCOphth Part 2 mock examination. PaLM 2 did not attain pass-worthy performance or match expert ophthalmologists’ scores but was within the spread of trainee doctors’ performance. LLMs are approaching human expert-level knowledge and reasoning in ophthalmology, and significantly exceed the ability of non-specialist clinicians (represented here by unspecialised junior doctors) to answer ophthalmology questions. Second, clinician grading of model outputs suggests that GPT-4 exhibits improved accuracy and relevance when compared with GPT-3.5. Development is producing models which generate better outputs to ophthalmological queries in the opinion of expert human clinicians, which suggests that models are becoming more capable of providing useful assistance in clinical settings. Third, LLM performance was consistent across question subjects and types, distributed similarly to human performance, and exhibited comparable agreement between other LLMs and doctors when corrected for differences in overall performance. Together, this indicates that the ophthalmological knowledge and reasoning capability of LLMs is general rather than limited to certain subspecialties or tasks. LLM-driven natural language processing seems to facilitate similar—although idiosyncratic—clinical knowledge and reasoning to human clinicians, with no obvious blind spots precluding clinical use.

Similarly dramatic improvements in the performance of GPT-4 relative to GPT-3.5 have been reported in the context of the North American Ophthalmology Knowledge Assessment Program (OKAP).^13,15^ State-of-the-art models exhibit far more clinical promise than their predecessors, and expectations and development should be tailored accordingly. Results from the OKAP also suggest that improvement in performance is due to GPT-4 being more well-rounded than GPT-3.5.^13^ This increases the scope for potential applications of LLMs in ophthalmology, as development is eliminating weaknesses rather than optimising in narrow domains. This study shows that well-rounded LLM performance compares well with expert ophthalmologists, providing clinically relevant evidence that LLMs may be used to provide medical advice and assistance. Further improvement is expected as multimodal foundation models, perhaps based on LLMs such as GPT-4, emerge and facilitate compatibility with image-rich ophthalmological data.^3,23,24^

### Limitations

This study was limited by three factors. First, examination performance is an unvalidated indicator of clinical aptitude. We sought to ameliorate this limitation by employing expert ophthalmologists, ophthalmology trainees, and unspecialised junior doctors answering the same questions as clinical benchmarks; and compared LLM performance to real cohorts of candidates in recent FRCOphth examinations. However, it remains an issue that comparable performance to clinical experts in an examination does not necessarily demonstrate that an LLM can communicate with patients and practitioners or contribute to clinical decision making accurately and safely. Early trials of LLM chatbots have suggested that LLM responses may be equivalent or even superior to human doctors in terms of accuracy and empathy, and experiments using complicated case studies suggest that LLMs operate well even outside typical presentations and more common medical conditions.^4,25,26^ In ophthalmology, GPT-3.5 and GPT-4 have been shown to be capable of providing precise and suitable triage decisions when queried with eye-related symptoms.^22,27^ Further work is now warranted in conventional clinical settings.

Second, while the study was sufficiently powered to detect a less than 10% difference in overall performance, the relatively small number of questions in certain categories used for stratification analysis may mask significant differences in performance. Testing LLMs and clinicians with more questions may help establish where LLMs exhibit greater or lesser ability in ophthalmology. Furthermore, researchers using different ways to categorise questions may be able to identify specific strengths and weaknesses of LLMs and doctors which could help guide design of clinical LLM interventions.

Finally, experimental tasks were ‘zero-shot’ in that LLMs were not provided with any examples of correctly answered questions before it was queried with FRCOphth questions from the textbook. This mode of interrogation entails the maximal level of difficulty for LLMs, so it is conceivable that the ophthalmological knowledge and reasoning encoded within these models is actually even greater than indicated by results here.^1^ Future research may seek to fine-tune LLMs by using more domain-specific text during pretraining and fine-tuning, or by providing examples of successfully completed tasks to further improve performance in that clinical task.^3^

### Future directions

Autonomous deployment of LLMs is currently precluded by inaccuracy and fact fabrication. Our study found that despite meeting expert standards, state-of-the-art LLMs such as GPT-4 do not match top-performing ophthalmologists.^28^ Moreover, there remain controversial ethical questions about what roles should and should not be assigned to inanimate AI models, and to what extent human clinicians must remain responsible for their patients.^3^ However, the remarkable performance of GPT-4 in ophthalmology examination questions suggests that LLMs may be able to provide useful input in clinical contexts, either to assist clinicians in their day-to-day work or with their education or preparation for examinations.^3,13,14,27^ GPT-4 may prove especially useful where access to ophthalmologists is limited: provision of advice, diagnosis, and management suggestions by a model with FRCOphth Part 2-level knowledge and reasoning ability is likely to be superior to non-specialist doctors and allied healthcare professionals working without support, as their exposure to and knowledge of eye care is limited.^27,29^

However, close monitoring is essential to avoid mistakes caused by inaccuracy or fact fabrication.^30^ Clinical applications would also benefit from an uncertainty indicator reducing the risk of erroneous decisions.^7^ As LLM performance often correlates with the frequency of query terms’ representation in the model’s training dataset, a simple indicator of ‘familiarity’ could be engineered by calculating the relative frequency of query term representation in the training data.^7,31^ Users could appraise familiarity to temper their confidence in answers provided by the LLM, perhaps reducing error. Moreover, ophthalmological applications require extensive validation, preferably with high quality randomised controlled trials to conclusively demonstrate benefit (or lack thereof) conferred to patients by LLM interventions. Trials should be pragmatic so as not to inflate effect sizes beyond what may generalise to patients once interventions are implemented at scale.^3,32,33^ In addition to patient outcomes, practitioner-related variables should also be considered: interventions aiming to improve efficiency should be specifically tested to ensure that they reduce rather than increase clinicians’ workload.^3^

## Conclusion

According to comparisons with expert and trainee doctors, state-of-the-art LLMs are approaching expert-level performance in advanced ophthalmology questions. GPT-4 attains pass-worthy performance in FRCOphth Part 2 questions and exceeds the scores of some expert ophthalmologists. As top-performing doctors exhibit superior scores, LLMs do not appear capable of replacing ophthalmologists, but state-of-the-art models could provide useful advice and assistance to non-specialists or patients where access to eye care professionals is limited.^27,28^ Further research is required to design LLM-based interventions which may improve eye health outcomes, validate interventions in clinical trials, and engineer governance structures to regulate LLM applications as they begin to be deployed in clinical settings.^34^

## Ethical approval

Ethical approval was not required for this study as human patients were not involved in any experiments.

## Data Availability

Data are available upon request, excluding copyrighted material from the textbook used for experiments.

## Abbreviations

AI: artificial intelligence
FRCOphth: Fellowship of the Royal College of Ophthalmologists
GPT: Generative Pretrained Transformer
LLaMA: Large Language Model Meta AI
LLM: large language model
OKAP: Ophthalmology Knowledge Assessment Program
PaLM: Pathways Language Model

## Ethical approval

Ethical approval was not required for this study, as human patients were not involved.

## Author contributions

AJT and DSJT conceived the study and coordinated the team. AJT, DSJT, and YLW contributed to study design. AJT, WF, RH, SM, and RS undertook LLM data collection. YLW, SWW, AM, YJC, SZ, AS, YHC, BKJT, NJ, and TFT answered questions independently to benchmark LLM performance. YLW, SWW, AM, YJC, and SZ appraised LLM outputs. AJT conducted data analysis and visualisation. AJT and DSJT drafted the manuscript. All authors participated in manuscript redrafting. DSJT provided academic supervision. All authors approved the submitted version of the manuscript.

## Acknowledgements

The authors extend their thanks to Mr Arunachalam Thirunavukarasu (Betsi Cadwaladr University Health Board) for his advice and assistance with recruitment.

